# Impact of population mask wearing on Covid-19 post lockdown

**DOI:** 10.1101/2020.04.13.20063529

**Authors:** Babak Javid, Nathalie Q. Balaban

**Affiliations:** Tsinghua University School of Medicine; Hebrew University of Jerusalem

## Abstract

COVID-19, caused by SARS-CoV2 is a rapidly spreading global pandemic. Although precise transmission routes and dynamics are unknown, SARS-CoV2 is thought primarily to spread via contagious respiratory droplets. Unlike with SARS-CoV, maximal viral shedding occurs in the early phase of illness, and this is supported by models that suggest 40-80% of transmission events occur from pre- and asymptomatic individuals. One widely-discussed strategy to limit transmission of SARS-CoV2, particularly from presymptomatic individuals, has been population-level wearing of masks. Modelling for pandemic influenza suggests some benefit in reducing total numbers infected with even 50% mask-use. COVID-19 has a higher hospitalization and mortality rate than influenza, and the impacts on these parameters, and critically, at what point in the pandemic trajectory mask-use might exert maximal benefit are completely unknown. We derived a simplified SIR model to investigate the effects of near-universal mask-use on COVID-19 assuming 8 or 16% mask efficacy. We decided to model, in particular, the impact of masks on numbers of critically-ill patients and cumulative mortality, since these are parameters that are likely to have the most severe consequences in the COVID-19 pandemic. Whereas mask use had a relatively minor benefit on critical-care and mortality rates when transmissibility (Reff) was high, the reduction on deaths was dramatic as the effective R approached 1, as might be expected after aggressive social-distancing measures such as wide-spread lockdowns. One major concern with COVID-19 is its potential to overwhelm healthcare infrastructures, even in resource-rich settings, with one third of hospitalized patients requiring critical-care. We incorporated this into our model, increasing death rates for when critical-care resources have been exhausted. Our simple model shows that modest efficacy of masks could avert substantial mortality in this scenario. Importantly, the effects on mortality became hyper-sensitive to mask-wearing as the effective R approaches 1, i.e. near the tipping point of when the infection trajectory is expected to revert to exponential growth, as would be expected after effective lockdown. Our model suggests that mask-wearing might exert maximal benefit as nations plan their post-lockdown strategies and suggests that mask-wearing should be included in further more sophisticated models of the current pandemic.

---

COVID-19, caused by SARS-CoV2 is a rapidly spreading global pandemic. Although precise transmission routes and dynamics are unknown, SARS-CoV2 is thought primarily to spread via contagious respiratory droplets^1^. Unlike with SARS-CoV, maximal viral shedding occurs in the early phase of illness ^1,2^, and this is supported by models that suggest 40-80% of transmission events occur from pre- and asymptomatic individuals^3,4^. One widely-discussed strategy to limit transmission of SARS-CoV2, particularly from presymptomatic individuals, has been population-level wearing of masks. Modelling for pandemic influenza suggests some benefit in reducing total numbers infected with even 50% mask-use^5^. COVID-19 has a higher hospitalization and mortality rate than influenza^6^, and the impacts on these parameters, and critically, at what point in the pandemic trajectory mask-use might exert maximal benefit are completely unknown.

We derived a simplified SIR model based on the population of Israel as proof of principle (population 8 million) to investigate the effects of near-universal mask-use on COVID-19 assuming 8 or 16% mask efficacy (see Methods for relevant parameters). We decided to model, in particular, the impact of masks on numbers of critically-ill patients and cumulative mortality, since these are parameters that are likely to have the most severe consequences in the COVID-19 pandemic. Whereas mask use had a relatively minor benefit on critical-care and mortality rates when transmissibility (R_eff_) was high (Fig. 1A), the reduction on deaths was dramatic as the effective R approached 1 (Fig. 1B), as might be expected after aggressive social-distancing measures such as wide-spread lockdowns^6^. One major concern with COVID-19 is its potential to overwhelm healthcare infrastructures, even in resource-rich settings, with one third of hospitalized patients requiring critical-care. We incorporated this into our model, increasing death rates for when critical-care resources have been exhausted, however, we also modelled the same parameters for scenarios in which critical care capacity was unrestricted (Fig. 1C-D). Our simple model shows that modest efficacy of masks could avert substantial mortality when critical care capacity is limiting, but also derives benefit when it is unrestricted. Importantly, the effects on mortality became hyper-sensitive to mask-wearing as the effective R approaches 1, i.e. near the tipping point of when the infection trajectory is expected to revert to exponential growth, as would be expected after effective lockdown.

**Figure 1.**
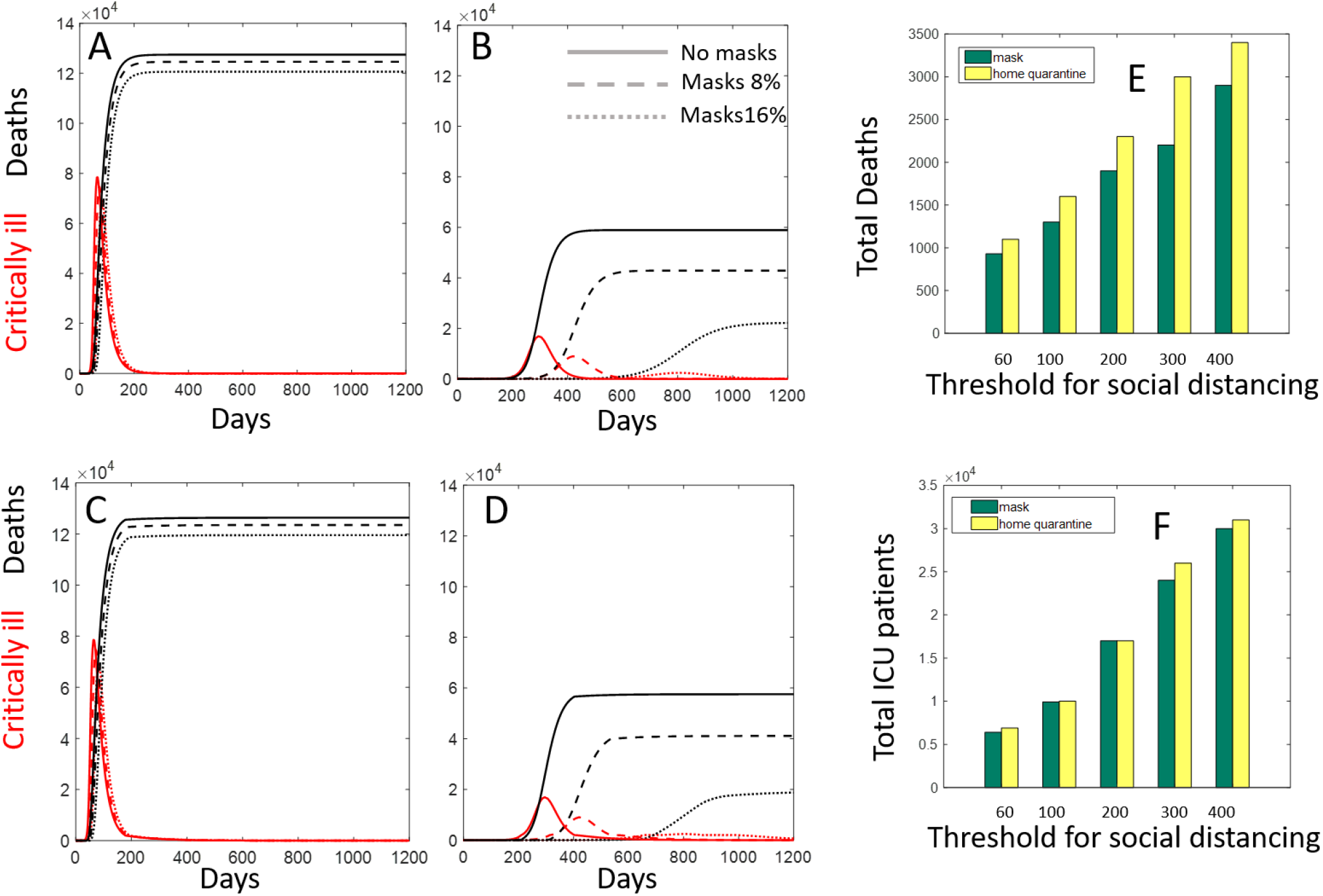
Mask effectiveness on mortality varies by R_eff_. (A) Number of critically ill patients (red) and total deaths (black) for an epidemic spreading with R of 2.2 (see Supplementary information for parameters) in a simple SIR model, x-axis represents time in days. The different curves are computed for a reduction of infectivity of 0, 8 and 16% by mask-wearing. (B) Same as A, but for an epidemic spreading with R of 1.3. Note that the reduction in infectivity by mask wearing has a larger effect. (C-D) Same as A-B but taking into account decrease in death when beds are unrestricted for critically-ill patients (see Supplementary information). (E-F) Analysis of the data of Ferguson *et al* (ref 5, Table 4). Assuming a 10% reduction in infectivity, mask wearing may be at least as effective as home confinement at reducing deaths (E) or preventing overwhelming icu beds (F). The different bars (1-5) are different thresholds (“triggers”) for implementing social measures in the Ferguson *et al* model.

In order to understand the generality of the effect of mask wearing upon home confinement removal, we also analysed the potential effects of mask-wearing for data provided by a more comprehensive and realistic model of the COVID-19 infection, which included modelling of different levels of social-distancing measures on infection and likely deaths^6^. When home-confinement is lifted but other social-distancing measures are in place, such as school closure and case isolation, wearing masks can maintain the benefits of home-confinement, both in terms of deaths (Fig. 1E) and critical-care bed use (Fig. 1F).

Limitations of our study include the relatively straightforward model we employed, as well as assumptions of high compliance with mask-wearing and their potential efficacy, for which definitive evidence in pandemics is lacking ^7,8^. Another recent modelling study of mask use came to similar conclusions as ours despite slightly different input parameters ^9^. However, that model mostly considered scenarios where the effective transmissibility of SARS-CoV2 remained high. Despite the limitations of our study, our model suggests that mask-wearing might exert maximal benefit as nations plan their ‘post-lockdown’ strategies and suggests that mask-wearing should be included in further more sophisticated models of the current pandemic. Since otherwise similar countries are currently devising different mask-wearing scenarios, the current situation offers an unprecedented opportunity to gather evidence on the real-world utility of population mask-wearing for implementation in this and future pandemics.

## Methods

### Infection dynamics model (Figure 1A-D)

To demonstrate the effect of masks, we used a simple SIR model of the dynamics of infection taking several populations into account: S: susceptible individuals, I: infected, R: resistant, CI: critically ill, D: dead. The goal of the model is not to predict any particular infection in a completely realistic way, but rather to illustrate the impact of reducing infectivity at high versus low R_0_ values.

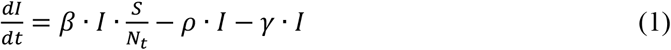

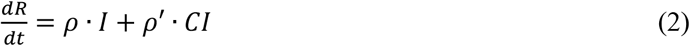

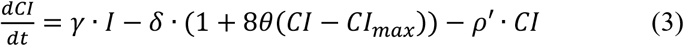

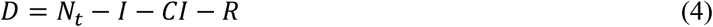

Where *θ*(*CI* − *CI_max_*) is the Heaviside function changing the death rate when the critically ill number saturates ICU beds. Parameters are defined in Table S1. Model was run using Matlab R2017a (MathsWorks,USA) ODE solver (‘ode23s’).

Wearing of masks was implemented in the model as a reduction of infectivity between 8-16% ^5,8,10–15^. Total population size was taken as 8×10^6^

| Parameter name | Symbol | Units | Values |
| --- | --- | --- | --- |
| Infection rate | $\beta$ | day<sup>-1</sup> | 0.17-0.4 |
| Recovery rate | $\rho$ | day<sup>-1</sup> | 0.16 |
| Critical deterioration rate | $\gamma$ | day<sup>-1</sup> | 0.003 |
| Death rate | $\delta$ | day<sup>-1</sup> | 0.0036 |
| Recovery of critically ill | $\rho'$ | day<sup>-1</sup> | 0.0025-0.005 |
| Total population size | $N_t$ | Individuals | $8 \times 10^6$ |
| Max. number of ICU beds | $CI_{max}$ | Beds | 2000 |

In the absence of ICU beds, 86% of the critical care patients die, whereas if ICU beds are not limiting, only 40% of critical care patients would die. The total fraction of critical care patients is 1.8% of the total number of infected cases^6^.

Data for (Figure 1E and 1F) was adapted from Ferguson *et al* ^6^ (16/3/2020-Table 4) The wearing of masks is assumed to reduce transmissibility by 10%. We, therefore, compared the results of Fergusson et al (Table 4) at different R and for different social-distancing policy measures.

Babak Javid MB BChir PhD

Tsinghua University School of Medicine

Nathalie Q. Balaban PhD

Racah Institute of Physics

The Hebrew University of Jerusalem

## Data Availability

All data are included in the manuscript and supplementary information, or will be made available by contacting the authors.

## Notes

### Competing Interest Statement

The authors have declared no competing interest.

### Funding Statement

No specific funding

